# Fixel-based analysis reveals macrostructural white matter changes associated with tau pathology in early stages of Alzheimer’s disease

**DOI:** 10.1101/2023.02.17.23286094

**Authors:** Khazar Ahmadi, Joana B. Pereira, Danielle van Westen, Ofer Pasternak, Fan Zhang, Markus Nilsson, Erik Stomrud, Nicola Spotorno, Oskar Hansson

**Affiliations:** Clinical Memory Research Unit, Department of Clinical Sciences, Lund University, Lund, Sweden; Institute of Cognitive Neuroscience, Faculty of Psychology, Ruhr University Bochum, Bochum, Germany; Division of Neuro, Department of Clinical Neurosciences, Karolinska Institutet, Stockholm, Sweden; Diagnostic Radiology, Department of Clinical Sciences, Lund University, Lund, Sweden; Department of Psychiatry, Brigham and Women’s Hospital, Harvard Medical School, Boston, Massachusetts, USA; Department of Radiology, Brigham and Women’s Hospital, Harvard Medical School, Boston, Massachusetts, USA; Department of Psychiatry, Massachusetts General Hospital, Harvard Medical School, Boston, Massachusetts, USA; Department of Medical Radiation Physics, Lund University, Lund, Sweden; Memory Clinic, Skåne University Hospital, Malmö, Sweden

**Keywords:** Alzheimer’s disease, diffusion tensor imaging, Fixel-based analysis, memory, white matter

## Abstract

White matter (WM) alterations are commonly found across different stages of Alzheimer’s disease (AD). However, the association between these changes with underlying AD pathology such as amyloid-β (Aβ) and tau deposition is still poorly understood. Hitherto, most studies have assessed WM alterations in AD using diffusion tensor imaging (DTI). Nonetheless, DTI has methodological shortcomings that limit an accurate biological interpretation. To address this limitation, here we applied fixel-based analysis (FBA) to disentangle microscopic differences in fiber density (FD) from macroscopic morphological changes in fiber cross-section (FC) in early stages of AD. We further investigated the associations of FBA metrics with AD pathology and cognitive performance. Additionally, we compared FBA results with other commonly used WM metrics derived from DTI, free-water corrected (FW)-DTI and diffusion kurtosis imaging (DKI). To achieve these goals, we included 224 Aβ-negative and 91 Aβ-positive cognitively unimpaired individuals as well as 78 Aβ-positive patients with mild cognitive impairment (MCI) with diffusion-weighted MRI (dMRI), Aβ-PET and tau-PET scans from the Swedish BioFINDER-2 study. We found that tau-PET uptake in medial temporal regions was associated with macrostructural alterations reflected by reduced FC mainly in the parahippocampal part of the cingulum bundle in Aβ-positive individuals. This tau-related WM alteration was also associated with impaired memory. Interestingly, only FBA metrics were able to capture the association between tau-PET uptake and white matter degeneration. No association was found between global amyloid load and any dMRI metrics. Compared to both cognitively unimpaired groups, MCI patients showed a decrease in all FBA metrics in the entire cingulum bundle, uncinate fasciculus and anterior thalamic radiations. Metrics derived from DKI, and FW-DTI revealed a similar pattern of alterations whereas the spatial extent of WM abnormalities detected by DTI was more widespread. Altogether, our results indicate that early WM alterations in AD are mainly due to macrostructural changes identified by FBA metrics, being more closely associated with tau than Aβ pathology. These findings suggest that future studies assessing the effects of AD pathology in white matter tracts should consider using FBA metrics.

## Introduction

Although Alzheimer’s disease (AD) is mainly regarded as a grey matter (GM) disease (Bejanin et al., 2017; Iaccarino et al., 2018; Möller et al., 2013), concomitant white matter (WM) degeneration has also been observed in postmortem studies (Brun & Englund, 1986; Englund, 1998). Such findings have been corroborated *in vivo* using diffusion tensor imaging (DTI) that has shown lower fractional anisotropy (FA) and higher mean diffusivity (MD) in several WM tracts in patients with mild cognitive impairment (MCI) and also in those with AD dementia (Liu et al., 2011; Mayo et al., 2017). Moreover, changes in DTI-derived metrics in familial AD have been reported to precede the onset of clinical symptoms (Araque Caballero et al., 2018; Li et al., 2015). However, the literature describing the link between WM changes and the underlying molecular pathology of AD is unclear. For instance, while the results of some studies based on DTI metrics suggest that increased diffusion restriction i.e., lower MD and higher FA is associated with amyloid-beta (Aβ) pathology in early stages of AD (Racine et al., 2014), other studies have shown an opposite association (Chao et al., 2013). Conversely, there are reports of no correlation between Aβ pathology and WM alterations (Kantarci et al., 2014; Strain et al., 2018).

Although these inconsistent findings may reflect cohort-specific differences, they might also arise from the methodological challenges associated with DTI. Due to partial volume effects, the metrics obtained with DTI are not specific to a single tissue type. This limitation is particularly relevant in studies on AD where vasogenic edema result in an increase in cerebrospinal fluid (CSF) and extracellular free water (FW) in the brain (Pasternak et al., 2009). This flaw can be mitigated using FW-DTI providing more accurate tissue-based metrics based on a bi-tensor model (Bergamino et al., 2021; Pasternak et al., 2009). Further, water diffusion in DTI is modeled based on the assumption that the displacement of water molecules has a Gaussian distribution while this assumption may not hold in biological tissues (Jensen & Helpern, 2010). The contribution of non-Gaussian diffusion can be quantified using Diffusion kurtosis imaging (DKI), which has been shown to be potentially more sensitive than DTI in detecting microstructural changes in WM (Gong et al., 2013; Wang et al., 2011; Zhang et al., 2021).

Another major limitation of DTI is the inability to resolve multiple fiber orientations in regions with crossing fibers that are present in up to 90% of the WM voxels (Jeurissen et al., 2013). This implies that differences in voxel-averaged metrics between groups may partially reflect differences in local multi-fiber geometry rather than microstructural differences. This could be the reason why some studies have found increased FA in subjects with mild cognitive impairment and mild AD (Douaud et al., 2011; Chad et al., 2018). The fixel-based analysis (FBA) framework is a higher-order diffusion model that enables characterization of multiple fiber orientations in a single voxel, facilitating fiber-tract-specific statistical comparisons (Raffelt et al., 2015, 2017). Further, FBA provides metrics estimated from fixels, that are distinct fiber populations within a voxel. These quantitative FBA metrics include fiber density (FD), which reflects microscopic changes in intra-axonal volume, fiber cross-section (FC), an index of macroscopic alterations in a cross-sectional area perpendicular to the WM bundles, and a combined measure which is a product of FD and FC i.e., FDC. In a previous study, this framework was used to examine WM degeneration in patients with MCI and AD dementia (Mito et al., 2018). The authors further studied the association of fiber-specific WM abnormalities with Aβ pathology. Their findings revealed WM alterations in tracts connecting key regions affected by AD. However, no association was found between FBA metrics and Aβ accumulation as quantified by PET. Moreover, in their study the association between WM changes and tau pathology was not investigated. The available studies on such associations are mainly based on DTI metrics and conducted on relatively small cohorts. Overall, these studies report an association between either higher MD or lower FA in the temporal WM tracts and tau-PET uptake in temporal regions (Carlson et al., 2021; Pereira et al., 2021; Strain et al., 2018). Only two recent studies have investigated the association of FBA metrics with tau pathology reporting partially inconsistent results (Luo et al., 2021; Dewenter et al., 2022). While the former work has shown an inverse relationship between FC in the ventral cingulum and increased tau-PET uptake in the entorhinal cortex (Luo et al., 2021), in the latter study tau-PET uptake was not associated to FBA metrics when accounting for the contribution of Aβ (Dewenter et al., 2022).

Given the above-mentioned inconsistent findings, we aimed to examine to what extent WM is compromised in early stages of the AD continuum and its association with both amyloid and tau pathologies as well as cognitive performance. To this end, we focused on a large cohort of non-demented individuals, including cognitively unimpaired participants and patients with MCI from the Swedish BioFINDER-2 study. Specifically, we investigated the difference between groups and assessed the relationship between WM changes and both Aβ- and tau-PET uptake. We applied a comprehensive set of approaches to parametrize the signal from diffusion MRI to identify WM alterations, including FBA, DTI, FW-DTI, and DKI.

## Methods

### Participants

This study comprised cognitively unimpaired individuals (CU) and MCI patients, recruited from the Swedish BioFINDER-2 study (<u>NCT03174938</u>) that has been previously described in detail (Palmqvist et al., 2020; Leuzy et al., 2020). The groups were further stratified into Aβ-negative or Aβ-positive based on the CSF Aβ42/40 ratio (Pichet Binette et al., 2022). Of 484 initially included participants, a total of 91 individuals were excluded due to excessive motion or other types of imaging artefacts (N=13), evidence of severe vascular co-pathology such as cerebral infarcts (N=10) and extensive white matter hyperintensities (WMH; N=68) that can affect the dMRI metrics (Svärd et al., 2017). This resulted in a final sample size of 393 individuals. A summary of the participants’ demographic and clinical characteristics is provided in Table 1. All participants gave written informed consent. The study procedures were in accordance with the Declaration of Helsinki and were approved by the Ethical Review Board in Lund, Sweden.

**Table 1.** Participants’ characteristics.

|  | **Aβ-negative CU**<br>**N = 224** | **Aβ-positive CU**<br>**N = 91** | **Aβ-positive MCI**<br>**N = 78** | **statistic** |
| --- | --- | --- | --- | --- |
| **Age** | 65.41<br>(9.74) | 69.97<br>(8.59) | 71.97<br>(6.44) | F = 19.26<br>p < 0.00001 |
| **Males/females** | 99/125 | 40/51 | 42/36 | $\chi^2 = 106.04$<br>p < 0.00001 |
| **APOE ε4**<br>**carriers%** | 35.26% | 70.32% | 75.64% | $\chi^2 = 155.6$<br>p < 0.00001 |
| **Years of**<br>**education** | 12.87<br>(3.59) | 12.48<br>(3.32) | 12.50<br>(4.45) | F = 0.507<br>p = 0.603 |
| **ADAS-cog**<br>**delayed word**<br>**recall** | 2.41<br>(1.71) | 3.17<br>(1.96) | 7.14<br>(2.20) | F = 184.7<br>p < 0.00001 |
| **VOSP-cube**<br>**analysis** | 9.63<br>(0.85) | 9.60<br>(1.12) | 8.59<br>(2.27) | F = 18.89<br>p < 0.00001 |
| **TMT (B-A)**<br>**[seconds]** | 44.19<br>(27.06) | 56.30<br>(52.14) | 139.39<br>(109.87) | F = 74.23<br>p < 0.00001 |
| **Global Aβ**<br>**(SUVR)** | 0.47<br>(0.04) | 0.65<br>(0.14) | 0.78<br>(0.17) | F = 248.4<br>p < 0.00001 |
| **Entorhinal tau**<br>**(SUVR)** | 1.09<br>(0.12) | 1.30<br>(0.26) | 1.58<br>(0.37) | F = 133.1<br>p < 0.00001 |

Data are presented as mean values followed by (standard deviations). Demographic and clinical variables were compared between groups using ANOVA or $\chi^2$ tests. CU = cognitively unimpaired, MCI = mild cognitive impairment, APOE $\epsilon$ 4 = Apolipoprotein E $\epsilon$ 4 allele, ADAS = Alzheimer's disease assessment scale-cognitive subscale, VOSP = visual object and space perception battery, TMT (B-A) = trail making test part B – trail making test part A, SUVR = standardized uptake value ratio. Note that a higher ADAS score indicates poor memory performance.

### Neuropsychological assessment

Different cognitive domains were assessed in the present study. Memory performance was measured using the ten-word delayed recall test from the Alzheimer’s Disease Assessment Scale-Cognitive Subscale (ADAS-Cog). Cube analysis from the Visual Object and Space Perception battery (VOSP) was used to examine visuospatial abilities. Finally, the difference in the scores of trail-Making Test (TMT) parts B and A was selected as a measure of executive function. Subtracting the time to complete part A from part B reduces visuospatial and working memory demands, hence providing a relatively pure indicator of the executive/attention performance (Tideman et al., 2022).

### MRI acquisition and processing

All participants underwent diffusion-weighted magnetic resonance imaging (dMRI) using a 3T MAGNETOM Prisma scanner with a 64-channel receiver coil array (Siemens Healthcare, Erlangen, Germany). The data were acquired using a single-shot EPI sequence with a multi-shell scheme using the following parameters: 2, 6, 32 and 64 gradient directions at b-values of 0, 100, 1000, and 2500 s/mm^2^, respectively, isotropic resolution of 2 mm^3^, phase-encoding direction = A-P, FOV = 220 × 220 × 124 mm^3^, multiband factor = 2, parallel imaging factor = 2, TR = 3500 ms, TE = 73 ms, and TA = 6:23 min. A second dMRI scan was also obtained with a reverse phase-encoding and 7 gradient directions (1 × b = 0 and 6 × b = 1000 s/mm^2^) for correction of susceptibility-induced distortions. Furthermore, a whole-brain T1-weighted scan (MPRAGE sequence, TR = 1900 ms, TE = 2.54 ms, voxel size = 1 × 1 × 1 mm^3^, FOV = 256 × 256 × 176 mm^2^, TA = 5:15 min) and a T2-weighted FLAIR scan (TR = 5000 ms, TE = 393 ms, TA = 4:37 min, same resolution and FOV as for the T1-weighted image) were acquired.

Preprocessing of dMRI data was performed using a combination of DIPY (https://dipy.org/; ‘*dipy_denoise_patch2self*’ and ‘*dipy_gibbs_ringing*’ functions), FSL (https://fsl.fmrib.ox.ac.uk/fsl/; ‘*topup*’ and ‘*eddy_openmp*’ with outlier replacement) and Mrtrix3 (www.mrtrix.org/; ‘*dwibiascorrect*’ command) software packages and included denoising, removal of Gibbs ringing artefacts, susceptibility off-resonance fields, motion/eddy current distortion correction and bias field correction (see also Garyfallidis et al., 2014; Andersson & Sotiropoulos, 2016; Andersson et al., 2003; Tournier et al., 2019). dMRI data were then quality controlled at a subject level using the FSL tool *‘eddy_quad’*. WMHs were automatically segmented in FLAIR images using the lesion prediction algorithm implemented in the lesion segmentation toolbox (https://www.applied-statistics.de/lst.html) of SPM. Moreover, total intracranial volumes were estimated as part of T1-weighted image processing in Freesurfer (https://surfer.nmr.mgh.harvard.edu/).

### FBA processing

FBA was performed according to the recommended pipeline of Mrtrix3 (Raffelt et al., 2017; Tournier et al., 2019). Briefly, tissue response functions for GM, WM and CSF were computed using ‘dhollander’ algorithm (Dhollander et al., 2016) based on which an average response function was obtained per tissue type across participants. Afterwards, dMRI data were upsampled to an isotropic voxel size of 1.3 mm^3^. Fiber orientation distributions (FOD) were estimated via the ‘Multi-Shell Multi-Tissue’ constrained spherical deconvolution (MSMT-CSD) using the group-averaged tissue response functions (Jeurissen et al., 2014). Subsequently, a multi-tissue informed log-domain intensity normalization was applied to achieve comparable FOD amplitudes between participants. Next, an unbiased center-specific WM FOD template was generated from 30 semi-randomly selected representative participants to which all individual FOD images were non-linearly registered (Raffelt et al., 2017). The resulting transformed FODs were segmented to create discrete fixels. FD was calculated as the integral of the warped FOD lobe corresponding to each fixel. FC was derived from the warp fields computed during the registration of individual FODs to the template space. The FC values were then log-transformed for downstream analysis to ensure that the data were centered around zero. A combined measure incorporating both the above metrics, i.e., FDC, was computed as the product of FD and FC (Dhollander et al., 2021). Finally, whole-brain probabilistic tractography was performed on the FOD template where initially 20 million streamlines were generated and subsequently filtered to 2 million streamlines using spherical-deconvolution informed-filtering of tractograms to reduce reconstruction bias (Smith et al., 2013). Connectivity-based fixel enhancement (CFE) using the template tractogram and nonparametric permutation testing with 5000 permutations were performed for statistical analysis (Raffelt et al., 2015).

### DTI, FW-DTI and DKI processing

To compare the results from FBA-derived metrics with more commonly used voxel-averaged measures, the preprocessed dMRI data were additionally fitted with tensor-derived models. Given that DTI and FW-DTI are traditionally applied to dMRI data with a lower number of gradient directions and b values, the 64 volumes with b = 2500 s/mm^2^ were not included in these analyses. DTI-derived metrics including FA and MD were quantified via the *‘dipy_fit_dti’* command of DiPY using weighted least-squares regression. FW-DTI measures i.e., FA_t_, MD_t_ and FW images were estimated with a bi-tensor model that has been described previously (Pasternak et al., 2009), implemented using an in-house MATLAB script. Since DKI requires multi-shell data, all volumes of the preprocessed dMRI data were included in this analysis. DKI parameters such as FA(DKI), MD(DKI) and mean kurtosis (MK) were calculated using the DIPY module *‘reconst*.*dki’* (Henriques et al., 2021). Afterwards, all individual tensor-derived maps were projected onto a standard space and skeletonized using Tract based spatial statistics (TBSS) toolbox in FSL (Smith et al., 2006). Voxel-wise analysis on the skeletonized tensor-derived metrics was conducted using FSL *‘randomise’* function with threshold-free cluster enhancement (TFCE) and 5000 permutations. An outline of the processing scheme for dMRI metrics is depicted in Figure 1.

**Figure 1.**
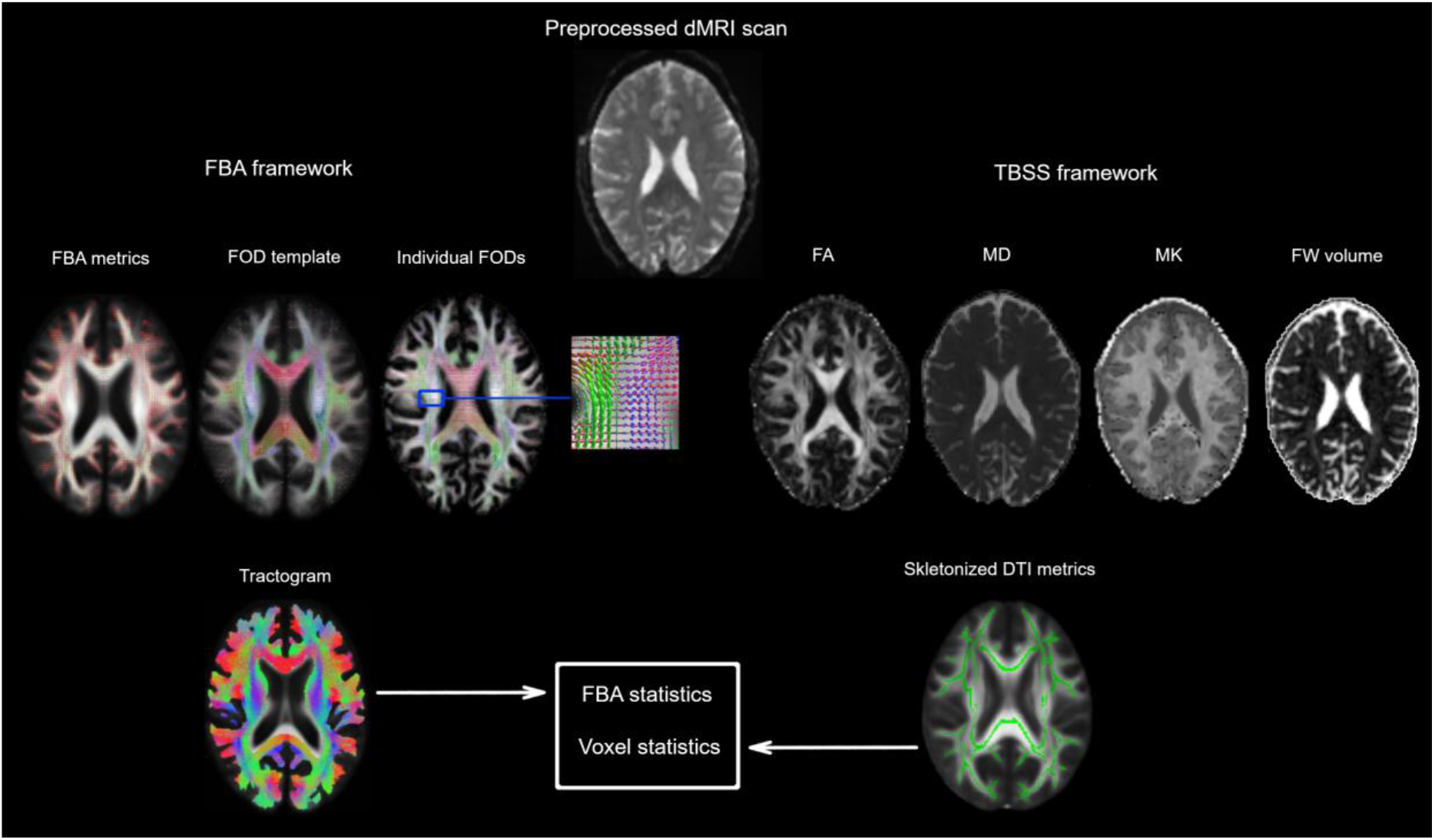
Overview of key processing steps of dMRI scans. Preprocessed dMRI data were used in FBA (left) and TBSS (right) frameworks. Note that for DTI and FW-DTI, volumes with high b-values were discarded from the preprocessed data. Individual FODs were obtained using MS-MT CSD and co-registered to a study-specific FOD template. Following estimation of FBA metrics (FD, FC, and FDC) for each warped FOD image, a whole-brain tractogram was generated to conduct FBA statistics using CFE. Tensor-derived metrics including FA, MD, MK and FW-volume were obtained using DTI, DKI and FW-DTI respectively. FA and MD images were also computed for DKI and FW-DTI but not displayed here. Voxel-wise analysis using TFCE was performed after co-registration of DTI-derived metrics to the template space and their subsequent skeletonization.

### PET acquisition and processing

Aβ positron emission tomography (PET) images were acquired on a GE Discovery MI scanner (General Electric Medical Systems) 90-110 minutes after the injection of [18F] flutemetamol. Tau-PET scans using [18F] RO948 were obtained on the same scanner 70-90 minutes post-injection as previously described (Leuzy et al., 2020). Standardized uptake value ratio (SUVR) images were calculated using the pons and inferior cerebellar GM as the reference regions for Aβ- and tau-PET, respectively (Pichet Binette et al., 2022; Leuzy et al., 2020). The acquired T1-weighted scans were used for PET-image co-registration and template normalization. Global Aβ burden was calculated using a neo-cortical composite region of interest (ROI) including prefrontal, parietal, temporal lateral regions as well as the anterior/posterior cingulate and precuneus (Landau et al., 2014; Lundqvist et al., 2013). Tau uptake was quantified primarily in the entorhinal cortex as one of the most representative regions for early tau accumulation. Nonetheless, other medial temporal lobe (MTL) regions i.e., hippocampus, amygdala and parahippocamups were also assessed for tau load. Note that Aβ- and tau-PET scans were not available in a small subsample of Aβ-negative CU (8 & 1), Aβ-positive CU (5 & 4) and Aβ-positive MCI participants (8 & 2) individuals, respectively.

### Statistical analysis

To explore the associations between both Aβ- and tau-PET uptake and dMRI-derived metrics, multiple univariate linear regression analyses were performed in the Aβ-positive individuals (Aβ-positive CU and Aβ-positive MCI). Similarly, between-group differences in dMRI-derived metrics were assessed using general linear models whose parameters were estimated via CFE or TFCE (see FBA, DTI, FW-DTI and DKI processing). The relationship between cognition and FBA-derived metrics was examined using ROI-based regression analyses performed in R (version 3.5.2) with each cognitive domain modelled as outcome measure and the FBA metrics as predictors. All models were controlled for the potential confounding effects of age, sex, ICV and WMH quantified by total lesion volume. Years of education was included as an additional nuisance covariate when testing the associations with cognitive performance. Moreover, the ‘mediation’ package in R was employed to assess whether the link between entorhinal tau load and memory performance was mediated by GM atrophy of this region or fiber-specific WM alterations in the parahippocampal part of the cingulum bundle. Statistical significance was set at family-wise error (FWE) corrected threshold of pFWE < 0.05 for both FBA and DTI-derived analyses.

## Data availability

Anonymized data will be shared by request from a qualified academic investigator for the sole purpose of replicating procedures and results presented in the article and as long as data transfer is in agreement with EU legislation on the general data protection regulation and decisions by the Ethical Review Board of Sweden and Region Skåne, which should be regulated in a material transfer agreement.

## Results

### 1. Association between dMRI-derived metrics and molecular pathology

#### Higher MTL tau load is associated only with decreased FC in the parahippocampal segment of the cingulum bundle

Elevated tau load in the entorhinal cortex was associated with lower FC almost exclusively in the bilateral parahippocampal portions of the cingulum bundle (Figure 2 A). The observed findings remained largely intact following additional adjustment for global Aβ load (Figure 2 B). A further sensitivity analysis including the GM volume of the entorhinal cortex in the model showed consistent, although partially less widespread results (see Figure 2 C). Similarly, increased tau-PET uptake in the other MTL regions was correlated with decreased FC mainly in the parahippocampal part of the cingulum, as illustrated in Supplementary Figure 1. Note that except for FC, no significant associations were found between other dMRI metrics and tau pathology. Aβ-PET uptake in the neo-cortical composite ROI was not correlated neither with FBA-nor DTI-derived measures.

**Figure 2.**
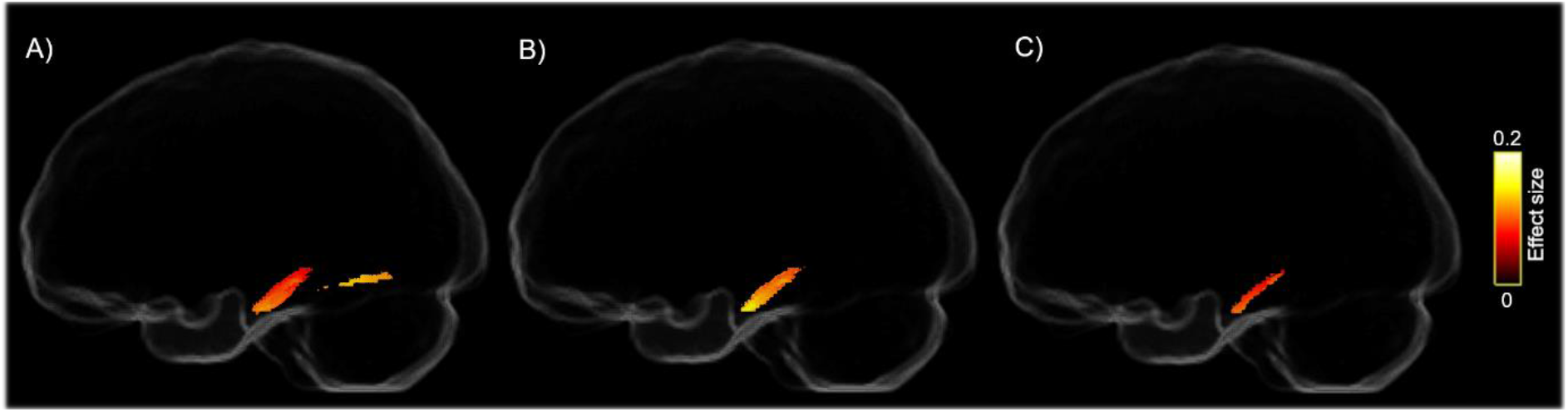
Negative association between entorhinal tau uptake and FC. A) higher levels of entorhinal tau are accompanied by lower FC in parahippocampal segment of the cingulum tract and inferior longitudinal fasciculus. The observed correlation remains significant although slightly decreased after controlling for B) global Aβ load and C) GM volume of the entorhinal cortex. Significant streamlines are displayed on the template glass brain (pFWE < 0.05).

### 2. Associations between WM alterations and cognitive performance

#### Decreased FBA metrics are correlated with memory decline

To test whether the tau-associated WM alterations were linked to worse cognitive functioning, mean values of each FBA metric were obtained for all Aβ-positive participants from a mask of fixels showing significant associations with the entorhinal tau load (see Figure 2 A). Worse memory performance correlated with a reduction in all three FBA metrics (Table 2). No relationship was found between other cognitive domains and FBA metrics (pFWE > 0.05) although a positive association between FC and visuospatial performance was close to the significance threshold (β = 0.22, uncorrected p = 0.039, pFWE = 0.11; see Table 2). Similar findings were obtained when assessing the correlation between cognition and fiber-specific WM abnormalities related to tau accumulation in other MTL regions (Supplementary Figure 2).

**Table 2.** Associations between tau-induced fiber-specific WM degeneration and different cognitive domains.

|  | **FD** | **FC** | **FDC** |
| --- | --- | --- | --- |
| **Memory performance** | $\beta = -0.25$<br>$p_{FWE} = 0.002$ | $\beta = -0.45$<br>$p_{FWE} = 3.9e-05$ | $\beta = -0.36$<br>$p_{FWE} = 1.9e-05$ |
| **Visuospatial abilities** | $\beta = 0.07$<br>$p_{FWE} = 1$ | $\beta = 0.22$<br>$p_{FWE} = 0.11$ | $\beta = 0.14$<br>$p_{FWE} = 0.31$ |
| **Executive functioning** | $\beta = -0.02$<br>$p_{FWE} = 1$ | $\beta = -0.005$<br>$p_{FWE} = 1$ | $\beta = -0.02$<br>$p_{FWE} = 1$ |

The regression models have been adjusted for age, sex, WMH, ICV and education. A decrease in FBA metrics parallels memory dysfunction.

Next, bootstrapped mediation analysis with 10000 iterations was performed to investigate possible effects of the GM volume of the entorhinal cortex and the observed tau-related WM alterations on the association between entorhinal tau accumulation and memory deficits. The GM volume of the entorhinal cortex partially mediated the tau-memory relationship (β = 0.83, CI = [0.–1 - 1.34], p < 0.0001, 20% meditation effect). However, fiber-specific WM alterations did not have a significant mediation effect (β = 0.23 | 0.48 | 0.47, CI = [-0.–3 - 0.67 | -0.21 – 1.28 | -0.23 – 1.13], p > 0.05 for FD, FC, and FDC, respectively. See also Supplementary Figure 3).

### 3. Group differences in FBA- and tensor-derived metrics

#### Reduced FBA metrics in MCI patients

Whole-brain FBA revealed significant decrease across all three FBA metrics in Aβ- positive MCI patients compared to Aβ-negative CU individuals (Figure 3 A). Lower FD was found in bilateral cingulum, parahippocampal parts of the cingulum bundle, uncinate fasciculi, anterior thalamic radiation and forceps minor. FC exhibited a similar pattern of results although primarily restricted to the left hemisphere. Analogous findings were observed for FDC with a larger effect size. Likewise, when the Aβ-positive MCI group was compared with Aβ-positive CU participants a similar, although more restricted, pattern of reduction in FBA metrics was found (Figure 3 B). In contrast, no group differences in any of the FBA-derived measures were found between Aβ-negative and Aβ-positive CU.

**Figure 3.**
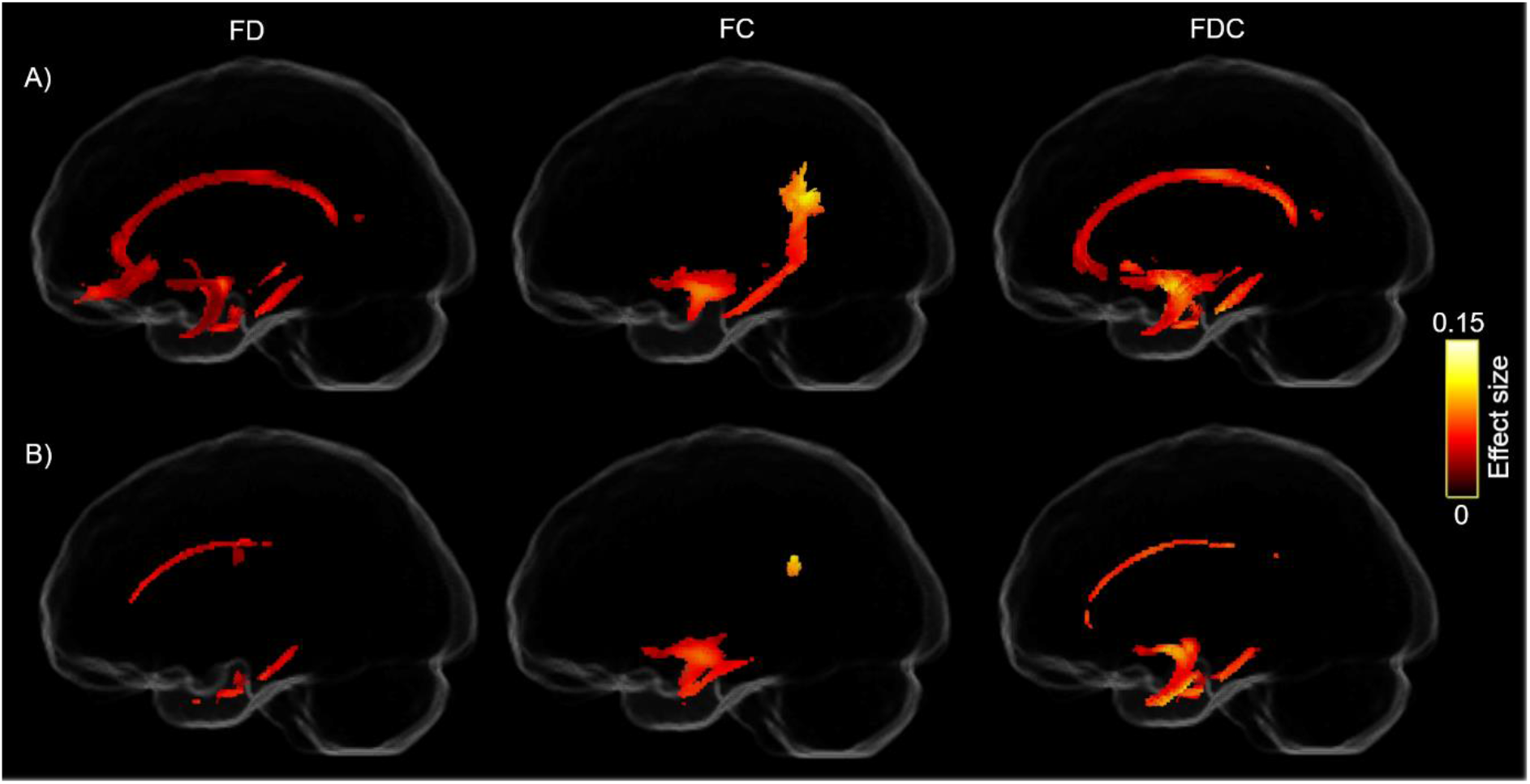
Comparison of FBA metrics between A) Aβ-negative CU and B) Aβ-positive CU individuals with Aβ-positive MCI patients. Streamlines were cropped from the template tractogram reflecting fixels with significantly reduced FD, FC and FDC (left, middle and right panels, respectively) in MCI patients. Significant streamlines are projected on the template glass brain (pFWE < 0.05).

#### Differences in tensor-derived metrics in MCI patients

Whole-brain voxel-wise analysis using TBSS demonstrated widespread decrease in FA and extensive increase in MD when comparing Aβ-positive MCI patients with both Aβ-negative and Aβ-positive CU individuals (Figure 4). Performing the same comparison using FW-DTI and DKI metrics reveled statistically significant differences only when employing FA_t_ and MK. Reduction in FA_t_ was found when comparing Aβ-positive MCI patients with both CU groups. However, MK reduction was observed only when the MCI patients were compared with Aβ-negative CU (see Figure 4). No significant differences between Aβ-negative and Aβ-positive CU individuals were found in any of the tensor-derived metrics.

**Figure 4.**
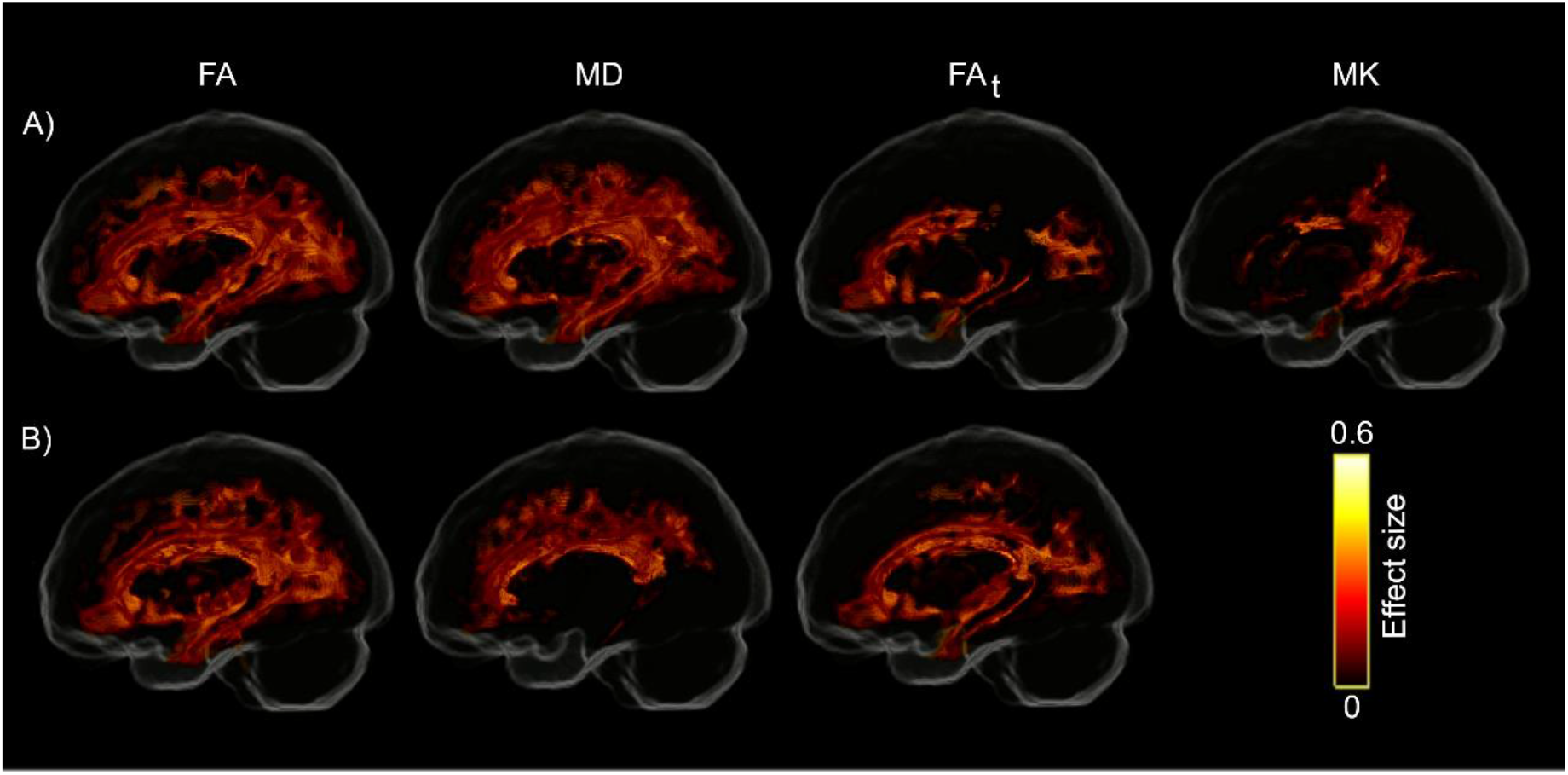
Voxel-wise comparison of tensor-derived metrics between A) Aβ-negative CU and B) Aβ-positive CU individuals with Aβ-positive MCI patients using TBSS. While FA, FA_t_ and MK are decreased, substantial increase of MD is observed in MCI patients (p_FWE_ < 0.05). WM tracts with significantly altered metrics are projected on the template glass brain.

## Discussion

Combining advanced dMRI acquisition and analytical techniques with PET imaging and neuropsychological measurements, we sought to investigate the involvement of WM degeneration in early stages of the AD pathological cascade. In Aβ-positive participants, elevated tau-PET uptake in medial temporal lobe structures was specifically associated with macroscopic changes of the parahippocampal part of the cingulum as indicated by lower FC. Moreover, the tau-related alterations in this WM bundle were correlated with a decline in memory performance. Importantly, the tau-WM association was not captured by the tensor-derived measures. This implies higher sensitivity of the FBA framework in detecting subtle WM degeneration, lending support to its utility as a potential biomarker for early detection and monitoring of tau-related AD progression. Using FBA, we further observed degeneration at both micro- and macroscopic scales mainly localized in cingulum, its parahippocampal segment, and frontal WM tracts in Aβ-positive MCI patients compared to Aβ-negative and Aβ-positive controls. Similar patterns were found using metrics derived from DTI, FW-DTI and DKI with a larger spatial extent for the DTI approach.

Notably, the parahippocampal portion of the cingulum connects key regions in the medial temporal and medial parietal lobes that are known to be affected during the early phase of AD pathogenesis (Zhou et al., 2008). The additional correction for global amyloid load did not alter the observed correlation, supporting an Aβ-independent association between tau accumulation and WM degeneration. However, further adjustments for the GM volume of the MTL regions decreased the effect size suggesting that the tau-FC association is not entirely independent from GM atrophy. Consistent with our findings, prior works have identified an association between increased tau-PET uptake and WM alterations in temporoparietal pathways (Binette et al., 2021; Luo et al., 2021; Jacobs et al., 2018; Strain et al., 2018).

The association of cognitive performance with FBA metrics in the same WM tract further supports the clinical relevance of our results indicating that the deterioration of memory functions is at least partially related to both micro- and macrostructural changes in WM. Further, mediation analysis was employed to investigate the relative contribution of GM and WM degeneration to memory impairment. Our results revealed that GM atrophy in the entorhinal cortex rather than WM alteration of the parahippocampal segment of the cingulum bundle mediated the relationship between elevated tau load in the entorhinal cortex and worse memory performance. These findings suggested that although memory deficits are linked to a synergetic degenerative process affecting both GM and WM, GM atrophy seems to bare a more direct impact on the association between tau accumulation and the deterioration of memory performance.

The lack of an association between amyloid accumulation and WM degeneration, as suggested by our results, is in agreement with previous reports using both DTI (Strain et al., 2018) and FBA (Mito et al., 2018). In contrast, other studies have shown either a negative or a positive correlation between FBA- or tensor-derived metrics with global Aβ-PET uptake in different stages of AD (Dewenter et al., 2022; Luo et al., 2021; Binette et al., 2021; Benitez et al., 2022). Although the observed discrepancy might arise from differences in the selection of participants or analytical approach, our results suggest that tau aggregation is more closely linked to WM alterations than Aβ pathology.

Whole-brain FBA analysis demonstrated diminished FD, FC and FDC in MCI patients in WM bundles associated with nodes of the default mode network connecting temporal, parietal and frontal cortices, supporting the network-based conceptualization of AD (Franzmeier et al., 2020; Mito et al., 2018). Remarkably, FD reduction was spatially more extensive than decrease in FC suggesting that microstructural changes of the affected WM tracts might be more pronounced than the macroscopical alterations of the same structures. Additionally, our FBA findings partially overlapped with between-group differences obtained with other dMRI metrics. FA_t_ and MK showed a spatially comparable pattern of results, whereas FA and MD revealed widespread abnormalities in the MCI patients affecting many WM bundles. The limitations of DTI in the accurate estimation of the metrics in regions with crossing fibers or the contamination of these metrics by extracellular free-water might result in inflated statistical results (Pasternak et al., 2009; Tournier et al., 2008). Nonetheless, the DTI-derived results of the current study are consistent with past evidence reporting WM alterations in early AD using DTI and its extensions (Metzler-Baddeley et al., 2014; Falangola et al., 2013; Gong et al., 2013; Teipel et al., 2010). By disentangling microstructural alterations from macroscopic WM changes, FBA provides fiber-specific insight, clarifying the present and previous DTI results that are likely driven by a combination of these differences.

Notably, no group differences were found in FBA- or DTI-derived metrics between Aβ-negative and Aβ-positive controls. While there is no study with direct comparison of FBA measures in those groups, conflicting results have been reported on group differences in DTI-derived metrics. Specifically, one study has shown increased MD in presymptomatic familial AD (Li et al., 2015) possibly due to more aggressive degeneration in genetic form of the disease. On the contrary, another study has found no significant differences in FA and MD between Aβ-positive and Aβ-negative cognitively unimpaired individuals (Fischer et al., 2015). Our findings are in line with the latter suggesting that amyloid burden per se does not induce WM degeneration.

The cross-sectional nature of this study did not allow us to determine the temporal relationship of WM alterations with PET uptake and cognitive measurements. Future studies are, thus, warranted to elucidate the longitudinal associations between WM degeneration, molecular pathology and clinical performance. Leveraging a large sample size, multi-shell dMRI data acquisition with high angular resolution, multiple diffusion models beyond DTI in conjunction with whole-brain fixel-based and voxel-wise analyses, the present study provides an exhaustive and robust framework for the evaluation of WM alterations and its association with the underlying pathology and core cognitive symptoms in the AD continuum. In conclusion, our results demonstrate that fiber-specific WM degeneration revealed by FBA is closely associated with tau accumulation and memory impairment even in early stages of AD.

## Authors Contribution

K.A: Conceptualization, Methodology, Formal analysis, Visualization, Writing – original draft; O.P: Methodology, Writing – review and editing; F.Z: Methodology, Writing – review and editing; D.W: Conceptualization, Writing – review and editing; J.B.P: Conceptualization, Writing – review and editing; M.N: Conceptualization, Methodology, Writing – review and editing; E.S: Data acquisition, Writing – review and editing; N.S: Conceptualization, Methodology, Writing – review and editing; O.H: Conceptualization, Supervision, Writing – review and editing.

## Supplementary material

**Supplementary Figure 1.**
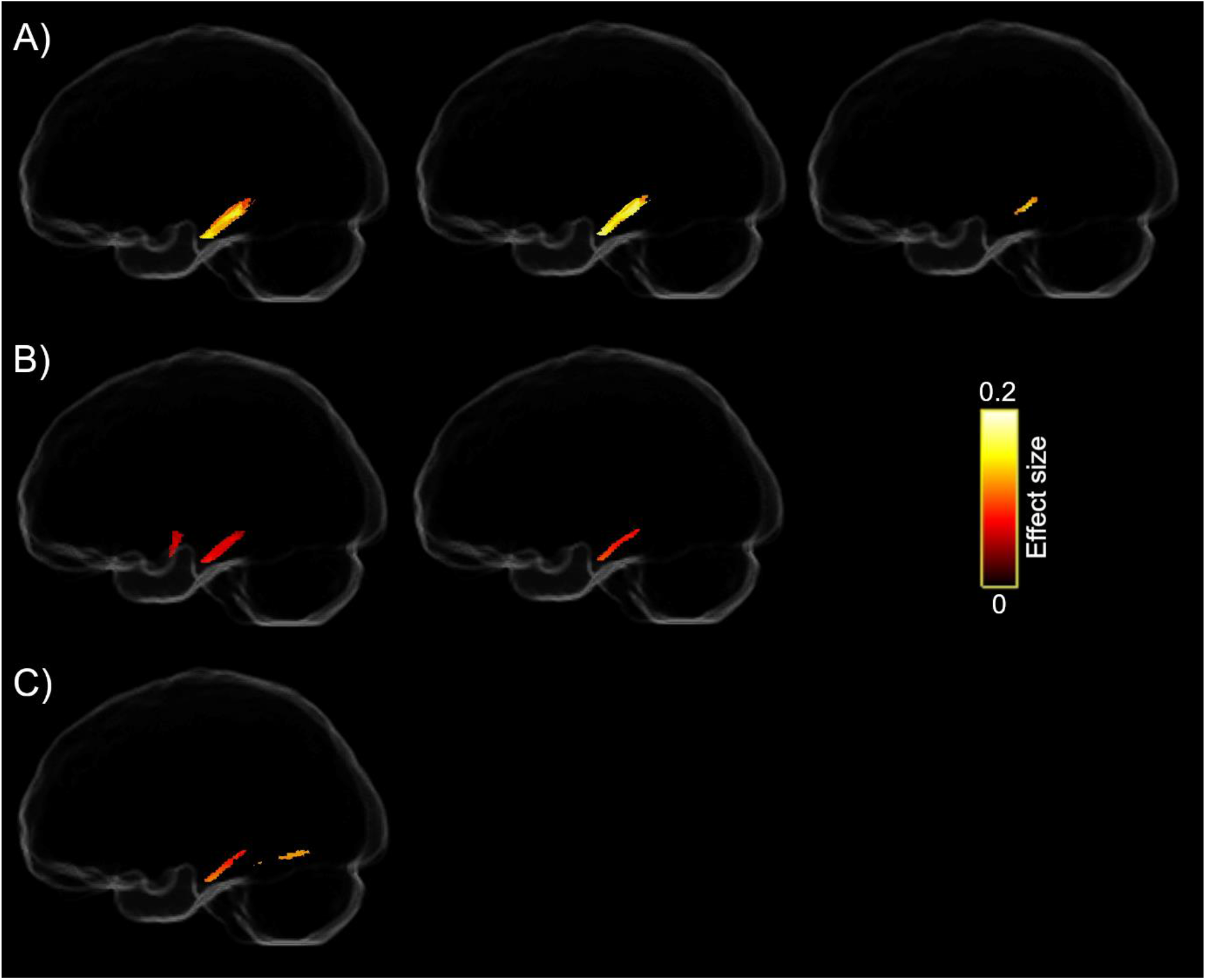
Association of FC with tau load in A) hippocampus, B) amygdala and C) parahippocampus. With increasing tau burden in medial temporal lobe regions, FC decreases primarily in the parahippocampal tract (pFWE < 0.05). Note that after controlling for global amyloid load (middle panel), the tau-FC association shrinks in amygdala and becomes insignificant in the parahippocampal cortex. Similarly, the observed correlations markedly decrease in the hippocampus and disappear in the other two regions following additional correction for the corresponding GM volume (right panel).

**Supplementary Figure 2.**
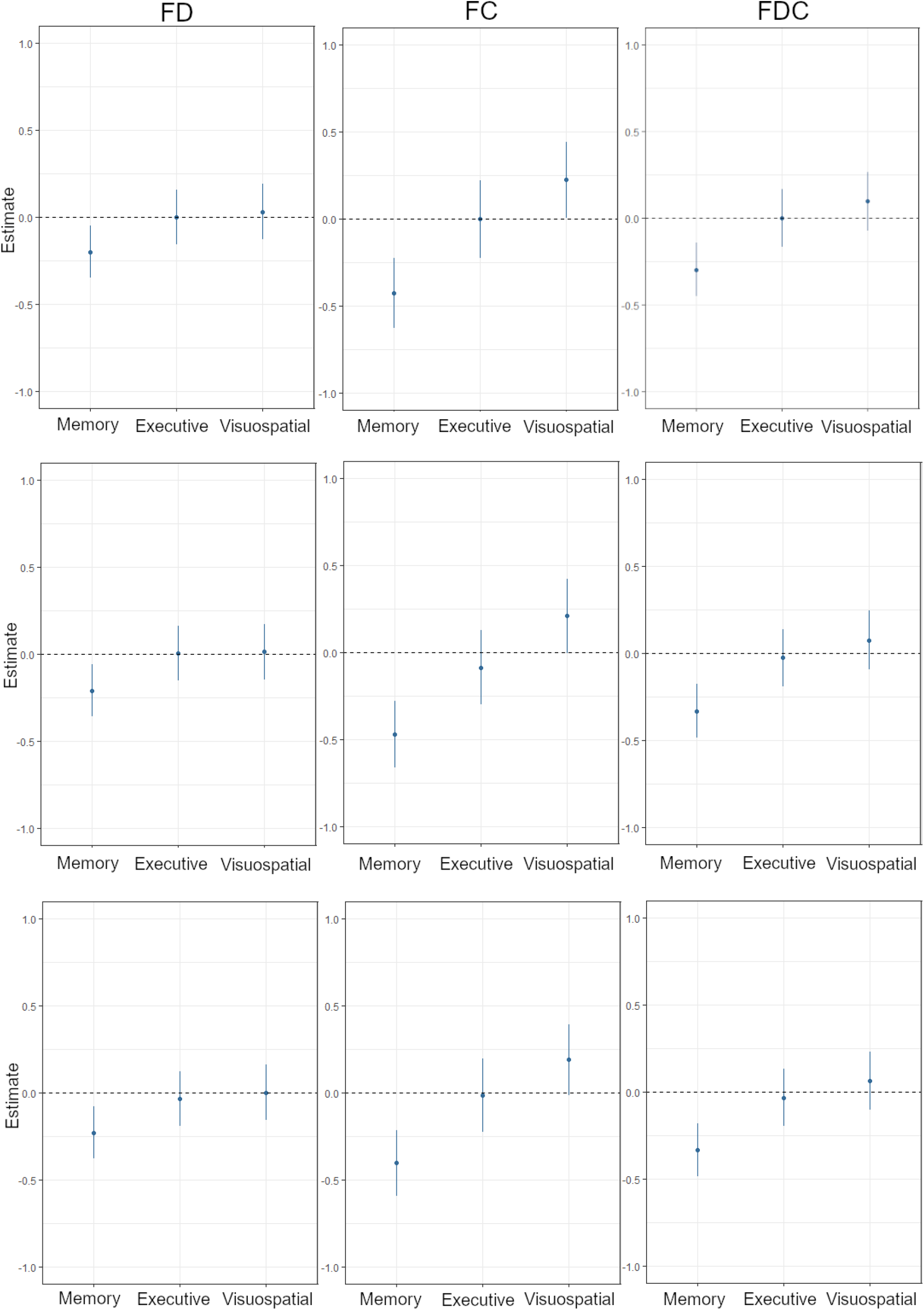
Regression coefficients (β) for the associations between different cognitive domains and fiber-specific WM degeneration in regions indicating significant correlations with tau load in the hippocampus, amygdala, and parahippocampus (top, middle and bottom panels, respectively; see Supplementary Figure 1). The regression models are corrected for the effects of age, sex, WMH, ICV and education. Lower FBA metrics are correlated with memory deficits (pFWE < 0.05).

**Supplementary Figure 3.**
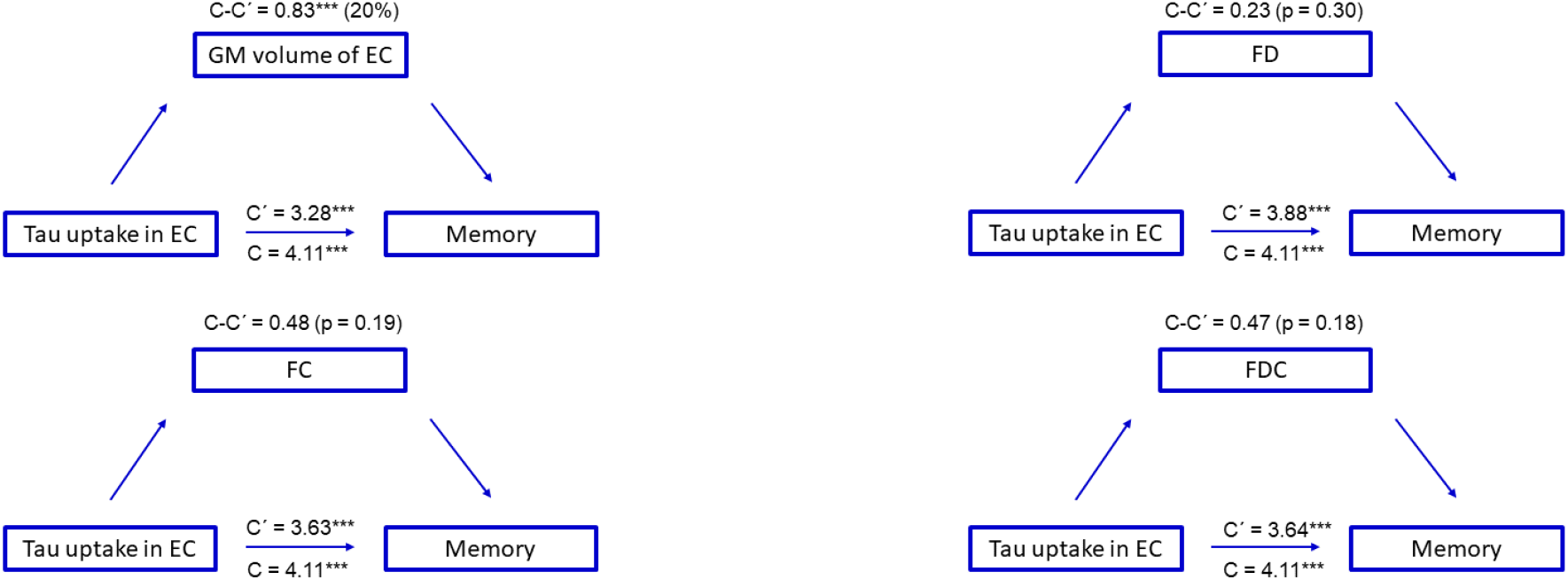
Flow chart illustration of the mediation analysis. The relationship between tau uptake in the entorhinal cortex (EC) is partially mediated by GM volume of this region (mediation effect = 20%) but no mediatory effect is observed for the fiber-specific WM alterations (top right and bottom panels). C presents the direct effect while C’ is the correlation strength after adjusting for mediatory variables. C-C’ is, therefore, the mediation effect.

